# Immunogenicity of BNT162b2 COVID-19 vaccine in New Zealand adults

**DOI:** 10.1101/2022.04.05.22273480

**Authors:** Frances H. Priddy, Michael Williams, Simon Carson, Brittany Lavender, Julia Mathieson, Chris Frampton, Nicole J. Moreland, Reuben McGregor, Georgia Williams, Maia Brewerton, Katie Gell, James Ussher, Graham Le Gros

## Abstract

**Background:** There is very little known about SARS-CoV-2 vaccine immune responses in New Zealand populations at greatest risk for serious COVID-19 disease.

**Methods:** This prospective cohort study assessed immunogenicity in BNT162b2 mRNA vaccine recipients in New Zealand without previous COVID-19, with enrichment for Māori, Pacific peoples, older adults ≥ 65 years of age, and those with co-morbidities. Serum samples were analysed at baseline and 28 days after second dose for presence of quantitative anti-S IgG by chemiluminescent microparticle immunoassay and for neutralizing capacity against Wuhan, Beta, Delta, and Omicron BA.1 strains using a surrogate viral neutralisation assay.

**Results:** 285 adults with median age of 52 years were included. 55% were female, 30% were Māori, 28% were Pacific peoples, and 26% were ≥65 years of age. Obesity, cardiac and pulmonary disease and diabetes were more common than in the general population. All participants received 2 doses of BNT162b2 vaccine. At 28 days after second vaccination, 99.6% seroconverted to the vaccine, and anti-S IgG and neutralising antibody levels were high across gender and ethnic groups. IgG and neutralising responses declined with age. Lower responses were associated with age ≥75 and diabetes, but not BMI. The ability to neutralise the Omicron BA.1 variant in vitro was severely diminished but maintained against other variants of concern.

**Conclusions:** Vaccine antibody responses to BNT162b2 were generally robust and consistent with international data in this COVID-19 naïve cohort with representation of key populations at risk for COVID-19 morbidity. Subsequent data on response to boosters, durability of responses and cellular immune responses should be assessed with attention to elderly adults and diabetics.

## INTRODUCTION

In 2019, a newly emergent coronavirus, Severe Acute Respiratory Syndrome coronavirus 2 (SARS-CoV-2) causing respiratory disease with potential for multi-system involvement (COVID-19) spread rapidly with significant morbidity, mortality and severe disruption of economic and social patterns worldwide. In New Zealand, early and stringent border control and a nationwide elimination policy limited SARS-CoV-2 spread during 2020 and most of 2021. Overseas, adults ≥65 years of age, elderly in aged care homes, those with co-morbidities such as cardiac and respiratory disease, diabetes and obesity were shown to be at higher risk of COVID-19-associated morbidity and mortality. This was mirrored in New Zealand, with Māori and Pacific peoples also being demonstrated to be at higher risk [1]. In March 2021, the BNT162b2 COVID-19 vaccine manufactured by Pfizer/BioNTech was offered to the New Zealand population. Little data on immunogenicity of COVID-19 vaccines is available in New Zealand populations. This prospective cohort study assessed immunogenicity in a subset of BNT162b2 recipients, with a focus on Māori, Pacific peoples, older adults ≥ 65 years of age, and those with co-morbidities.

There are several reasons to evaluate immune responses in local populations. The pivotal efficacy trials for COVID-19 vaccines enrolled minimal numbers of Pacific populations and did not include Māori. While vaccines are generally believed to act similarly across populations, reduced immunogenicity and effectiveness have been demonstrated for specific vaccines in the elderly due to immune senescence, and populations in low-resource settings possibly due to increased immune activation [2]. Māori and Pacific populations in New Zealand have disproportionate rates of cardiac and pulmonary disease, diabetes and obesity which are likely to increase morbidity from COVID-19. Modeling of COVID-19 cases during 2020 and early 2021 showed that Māori had 2.5 times greater odds of hospitalization than non-Māori, non-Pacific peoples, while Pacific peoples had 3 times greater odds [1]. During the period of Delta and Omicron transmission from August 2021 to the present, Māori accounted for 21% of cases and 25% of hospitalised cases. Pacific peoples accounted for 16% of cases and 34% of hospitalized cases [3]. Older adults ≥50 years of age accounted for only 18% of cases during this period, but 47% of hospitalizations [3]. Confirming immunogenicity profiles in these populations provides important data to inform the national COVID-19 strategy and to enhance vaccine confidence.

Pivotal efficacy trials were conducted in regions with high levels of SARS-CoV-2 circulation and transmission, which differs significantly from New Zealand which had limited exposure and community spread prior to 2022. Vaccine immune responses and the need for boosting may differ in populations with minimal prior viral exposure. Since the beginning of the pandemic, several variants of SARS-CoV-2 with increased transmissibility have evolved, with community transmission of Delta variant beginning in August 2021 and of Omicron variant in Jan 2022 in New Zealand. The BNT162b2 COVID-19 vaccine currently in use is based on sequences from the ancestral (Wuhan) strain and has demonstrated variable ability to neutralise variants of concern (VoC) in vitro, with sustained neutralisation against Delta, a modest decrease against Beta and significant decrease against Omicron [4,5]. These changes in neutralisation capacity correspond to similar patterns in decreased efficacy against infection across VoCs, while protection against severe disease is mildly reduced for Delta, with a more significant reduction for Omicron [6,7].

It is important to be able to evaluate the ability of vaccine-induced immune responses in the population to neutralise current or future variants. Well-characterized serologic specimens from vaccine recipients can also be used to evaluate the use of new, booster vaccines for COVID-19 variants, and to identify serologic tests of vaccine immunity for use in occupational health or travel. In addition, COVID-19 vaccination across a diverse population provides an important opportunity to evaluate host factors that may influence the effectiveness of vaccines. There are several lines of evidence suggesting a relationship between host factors such as immune activation and gut microbiome and protective antibody responses to vaccination [8].

The ongoing Ka Mātau, Ka Ora study is assessing immunogenicity in a subset of BNT162b2 COVID-19 vaccine recipients, with a focus on Māori, Pacific peoples, older adults ≥ 65 years, and those with co-morbidities. Vaccine immune responses were characterized and compared to international data and proposed correlates of immunity. The ability of vaccine-induced responses to neutralise new viral variants was evaluated.

## METHODS

### Study design and population

Participants were recruited from people eligible for COVID-19 vaccination in New Zealand from 10 June 2021 to 18 September 2021 at two study centers. The study sites were selected to access population groups across both the North and South Islands of New Zealand, to avoid the previous small COVID-19 outbreak center in the far south of the country, and to avoid study disruption from potential future outbreaks in the main population center of Auckland. During the majority of the enrollment period, no widespread community transmission of SARS-CoV-2 was detected in the country. From 17 August 2021 community transmission of the Delta variant was detected in the country but was not detected around the study centers until after the enrollment period ended.

Because the New Zealand vaccination program was sequenced according to occupation and risk, the populations eligible for vaccination according to national guidelines during the study enrollment period included (1) at-risk people age ≥16 living in settings with a high risk of exposure to COVID-19, (2) people age ≥65 and (3) people age ≥16 with underlying health conditions or disabilities. Relevant underlying health conditions included coronary heart disease, hypertension, stroke, diabetes, chronic obstructive pulmonary disease/chronic respiratory conditions, kidney disease, cancer and pregnancy. The Pfizer/BioNTech BNT162b2 COVID-19 vaccine was the preferred vaccine for use in NZ during this period.

Key inclusion criteria for the study were age ≥16 and eligibility for COVID-19 vaccination by national guidelines. Participants were excluded if they had a history of COVID-19, current respiratory or constitutional symptoms, recent international travel, systemic immunosuppressant medications, recent receipt of blood products or were a close contact of a confirmed or suspected COVID-19 case. Enrollment was enriched to ensure adequate representation of Māori, Pacific peoples, older adults ≥65 and women. Although pregnancy was not an exclusion criterion, no pregnant women were included in the study as COVID-19 vaccination was not widespread for pregnant women during this period when community transmission was very low.

All participants provided written informed consent. Demographic, past medical history, medications and BMI were recorded at baseline. Ethnicities were collected using NZ census categories. Selected co-morbidities associated with increased risk for COVID-19 morbidity and mortality were collected including overweight and obesity, cardiac disease (heart failure, coronary artery disease, cardiomyopathy and hypertension), diabetes (type 1 and 2), and pulmonary disease (chronic obstructive pulmonary disease, moderate to severe asthma, smoking, vaping)

Blood samples were taken within 7 days prior to first vaccination, within 3 days prior to second vaccination, and 28 days after second vaccination, with an allowable window from 21 to 56 days after second vaccination. All vaccinations were conducted by the national COVID-19 vaccination program, using the Pfizer/BioNTech BNT162b2 COVID-19 vaccine. The standard 30ug dose was given by intramuscular injection approximately 21 days apart. Data on needle length or site of injection was not collected, however both 25 mm and 28 mm needles were made available in the national vaccination program. Vaccination status and date of vaccination were determined by self-report and review of participants’ vaccination cards. Reactogenicity and other safety information were not collected as part of this study. In September 2021, NZ recommended a longer interval of 6 weeks between first and second dose, however the majority of study participants were able to maintain the original approximately 21 day interval.

### Laboratory analysis

#### Anti-SARS-CoV-2 Nucleocapsid IgG

Serum samples were analysed by Southern Community Laboratories in Dunedin NZ with the Abbott SARS-CoV-2 IgG assay according to the manufacturers’ instructions (Abbott Laboratories, Abbott Park USA). The assay is a chemiluminescent microparticle immunoassay designed to detect IgG antibodies to the nucleocapsid (NC) protein of SARS-CoV-2 in serum and plasma from individuals who are suspected to have had COVID-19. The cut-off value is 1.4 Index. Assay specificity was calculated locally as 99.6% using antenatal samples (n=300) in New Zealand [9].

#### Anti-SARS CoV-2 Spike IgG

Serum samples were analysed by Southern Community Laboratories in Dunedin NZ with the Abbott SARS-CoV-2 IgG II Quant assay according to the manufacturers’ instructions (Abbott Laboratories, Abbott Park USA). The assay is a chemiluminescent microparticle immunoassay for the quantitative detection of IgG antibodies to the RBD of the spike (S) protein with sensitivity and specificity of 99.37% and 99.55% respectively according to the manufacturers data. Assay specificity was calculated locally as 100% using antenatal samples (n=100) in New Zealand [10]. The cut-off value is 50 AU/mL. This assay has been calibrated by the manufacturer using the WHO international standard for SARS-CoV-2. AU/mL results were converted into IU/mL using a conversion factor supplied by the manufacturer.

#### SARS-CoV-2 neutralising antibodies

Serum samples were analysed at University of Auckland using a SARS-CoV-2 Surrogate Virus Neutralisation Test (sVNT) marketed as cPASS (GenScript, Singapore). This assay measures neutralising antibodies by quantifying the antibodies that block binding of the SARS-CoV-2 receptor binding domain to the hACE-receptor. Specificity was previously determined to be 100% using 413 local samples [11]. Sera was diluted 1:10 and the assay was performed following published protocols [12,13]. Samples were considered positive if percent inhibition was above 20% as previously defined. Pre-vaccine samples were initially assessed against the ancestral strain to determine if any of the cohort had SARS-CoV-2 neutralising antibodies pre-vaccine indicative of prior infection. Post-vaccination samples were assessed against the ancestral strain as well as to the Beta, Delta and Omicron BA.1 variants of concern (VoC). This assay has been calibrated using the WHO international standard for SARS-CoV-2. Percent inhibition results were converted to IU/mL titers using an online tool validated for the ancestral strain [14]. The conversion was not performed for the VoCs.

### Statistical analysis

Statistical analyses were performed using SPSS v27 software. SARS-CoV-2 IgG titers are summarized as geometric means with 95% confidence intervals. SARS-CoV-2 neutralising antibody responses are expressed as percentage inhibition, These responses were log_e_ transformed and summarized as back-transformed percentage inhibition means with 95% confidence intervals. The mean levels are compared between demographic groups defined by ethnicity, age, BMI, gender and individual and combined comorbidity status using factorial ANOVA of log_e_ transformed values. Where the ANOVA indicates significant effects these were further explored within factors using Fisher’s protected least significant difference test, and summarised as geometric mean ratios with 95% confidence intervals. Pearson’s correlation coefficients (r) were used to determine the correlation between titers obtained by the SARS-CoV-2 IgG anti-S assay and the surrogate viral neutralisation assay. Comorbidity prevalence was compared between ethnicity groups using logistic regression analyses adjusting for age and BMI groups. Statistical significance was set at p<0.05. Graphical representation of the data was performed using R (version 4.0.3) and RStudio (version 1.3.1093).

### Ethical approval and Māori consultation

The study was approved by the NZ Ministry of Health COVID-19 Emergency Response Health and Disability Ethics Committee (21/NTB/117). Māori consultation was provided by representatives in the study site communities, and Te Urungi Māori advisory group of the Malaghan Institute of Medical Research.

## RESULTS

This report contains primary immunology findings through 28 days after second vaccination for all participants. From 10 June 2021 to 18 September 2021, 331 individuals were screened and 298 enrolled into the study (Figure 1). 13 participants (4.4%) withdrew or were excluded from analysis prior to the primary immunology timepoint at Visit 3, 28 days after the second vaccination (Figure 1). In the 285 participants analysed, there were no reports of previous COVID-19 infection. Study participant age ranged from 16-92 years (median 52 years, IQR 36-66) and 156 (55%) were female. For analysis of ethnicity the NZ census category of European was used and includes NZ European as well as other Europeans. The proportions of Māori and Pacific peoples were higher than the general population according to the 2018 NZ census, 30.2% v. 16.2% for Māori, and 28.1% v. 6.6% for Pacific peoples (Table 1). The European subgroup had a higher median age (62 years) than Māori (49.5 years) or Pacific peoples (42.5 years). Overweight and obesity were highly prevalent among the study population (84%) which was also characterized by relatively high prevalence of other comorbidities associated with increased COVID-19 morbidity. After controlling for age differences among ethnic subgroups, there were statistically significant differences in the prevalence of overweight, obesity, cardiac disease, pulmonary disease and diabetes among ethnic subgroups. The study did not target immunocompromised populations and excluded participants on immune suppressive or immunomodulatory medication.

**Figure 1.**
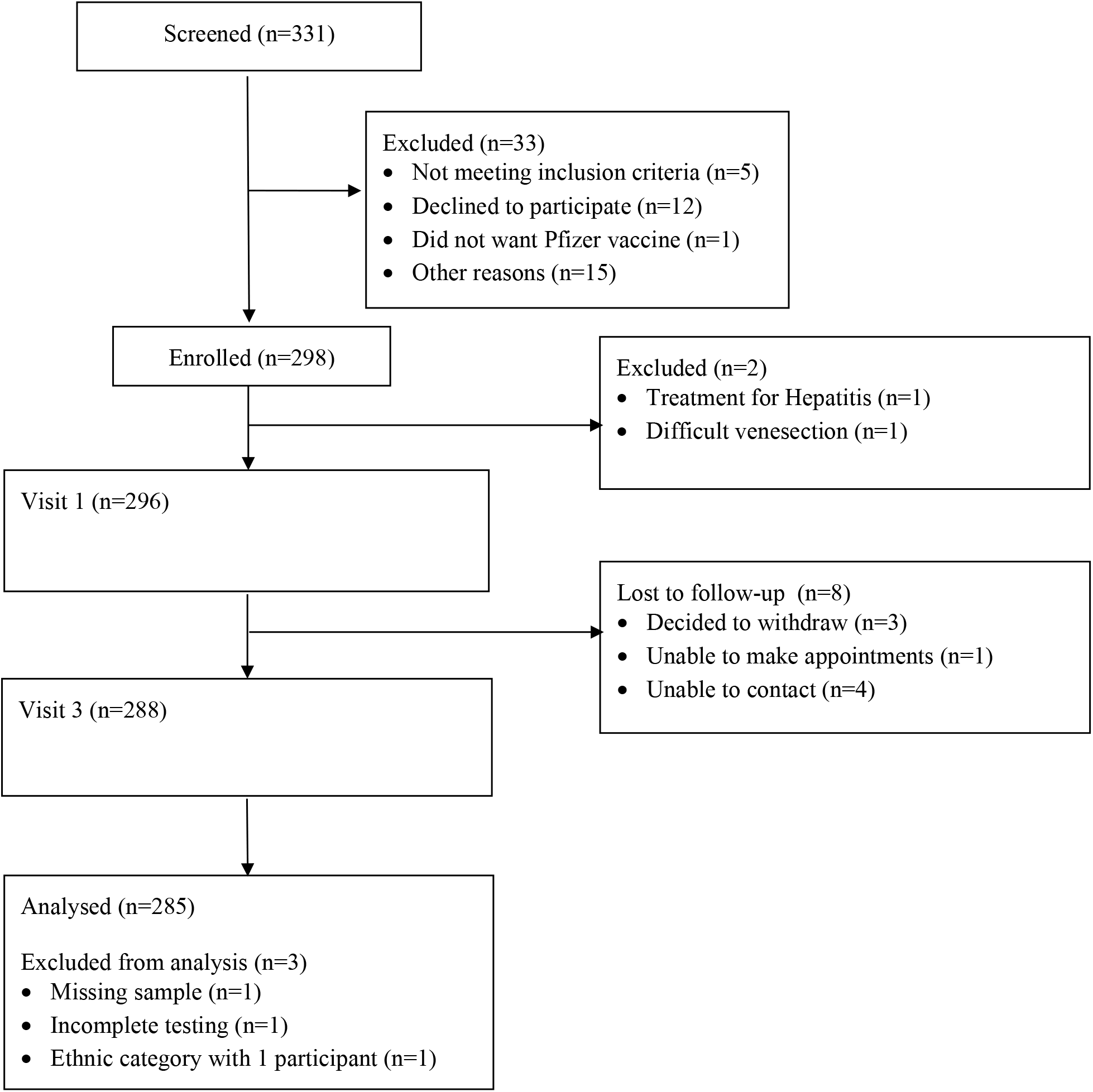
Participant flow through the study

**Table 1.** Study participant demographics

|  | % (n) | Median age<br>(IQR) | Comorbidities |  |  |  |  |  |
| --- | --- | --- | --- | --- | --- | --- | --- | --- |
|  |  |  | Obesity<br>% (n) | Overweight<br>% (n) | Cardiac<br>% (n) | Hypertension<br>% (n) | Diabetes<br>% (n) | Pulmonary<br>% (n) |
| **All Participants** | 285 | 52 (36-66) | 56.1 (160) | 28.1 (80) | 20.4 (58) | 18.6 (53) | 9.8 (28) | 16.1 (46) |
| **Gender** |  |  |  |  |  |  |  |  |
| Female | 54.7 (156) | 50 (34-65) | 55.1 (86) | 28.8 (45) | 20.5 (32) | 19.2 (30) | 9.6 (15) | 21.2 (33) |
| Male | 44.9 (128) | 55 (39-66) | 57.0 (73) | 27.3 (35) | 20.3 (26) | 18.0 (23) | 10.2 (13) | 10.2 (13) |
| Non-binary | 0.4 (1) | 31 | 100.0 (1) | 0.0 (0) | 0.0 (0) | 0.0 (0) | 0.0 (0) | 0.0 (0) |
| **Age** |  |  |  |  |  |  |  |  |
| 16-34 | 24.6 (70) | - | 49.0 (35) | 21.4 (15) | 2.9 (2) | 2.9 (2) | 1.4 (1) | 11.4 (8) |
| 35-54 | 30.5 (87) | - | 65.5 (57) | 31.0 (26) | 11.5 (10) | 11.5 (10) | 12.6 (11) | 21.8 (19) |
| ≥55 | 44.9 (128) | - | 53.1 (68) | 30.5 (39) | 35.9 (46) | 32.0 (41) | 12.5 (16) | 14.8 (19) |
| ≥65 | 25.6 (73) | - | 52.1 (38) | 27.4 (20) | 49.3 (36) | 43.8 (32) | 15.1 (11) | 15.1 (11) |
| **Ethnicity** |  |  |  |  |  |  |  |  |
| European | 41.8 (119) | 62.0 (49-76) | 41.2 (49) | 33.6 (40) | 26.9 (32) | 24.4 (29) | 5.9 (7) | 13.5 (16) |
| Māori | 30.2 (86) | 49.5 (35.3-59)** | 60.5 (52)** | 30.2 (26) | 16.3 (14)* | 14.0 (12) | 4.7 (4) | 25.6 (22)* |
| Pacific peoples | 28.1 (80) | 42.5 (25.8-52.5)** † | 73.8 (59)** | 17.5 (14)* | 15.0 (12)* | 15.0 (12) | 21.3 (17)** | 10.0 (8)** |

\*p<0.05 v European
\*\*p<0.01 v European
†p<0.05 v Māori
Obesity BMI ≥30 kg/m<sup>2</sup>; Overweight BMI >25 kg/m<sup>2</sup> and <30 kg/m<sup>2</sup>
Cardiac includes heart failure, coronary artery disease, cardiomyopathy, hypertension.
Diabetes is primarily type 2 with a single case of type 1
Pulmonary includes chronic obstructive pulmonary disease, moderate to severe asthma and smoking or vaping
Pacific peoples includes Samoan (45), Tongan (11), Tokelauan (9), Cook Island Māori (7), Fijian (5), Other (3)

All participants received two doses of BNT162b2. Approximately 73% of participants maintained the 21 day interval. The shortest interval was 15 days and the longest interval was 88 days between first and second dose.

### Seropositivity after vaccination

At baseline prior to vaccination, anti-NC IgG was negative in all participants indicating no current or prior COVID-19 infection (data not shown). All but one participant had negative anti-S IgG levels at baseline (Figure 2a). The participant had negative anti-NC IgG at baseline, no history of known COVID-19 infection, infected contacts or vaccination. At 28 days after second vaccination, 284 (99.6%) of participants had detectable Anti-S IgG (Figure 2a). The only non-responder was >75 years of age with a chronic hematologic cancer, treated with steroids. This participant subsequently received a third primary vaccine dose as well as a booster (4^th^ dose) consistent with national guidelines for immunocompromised people.

**Figure 2.**
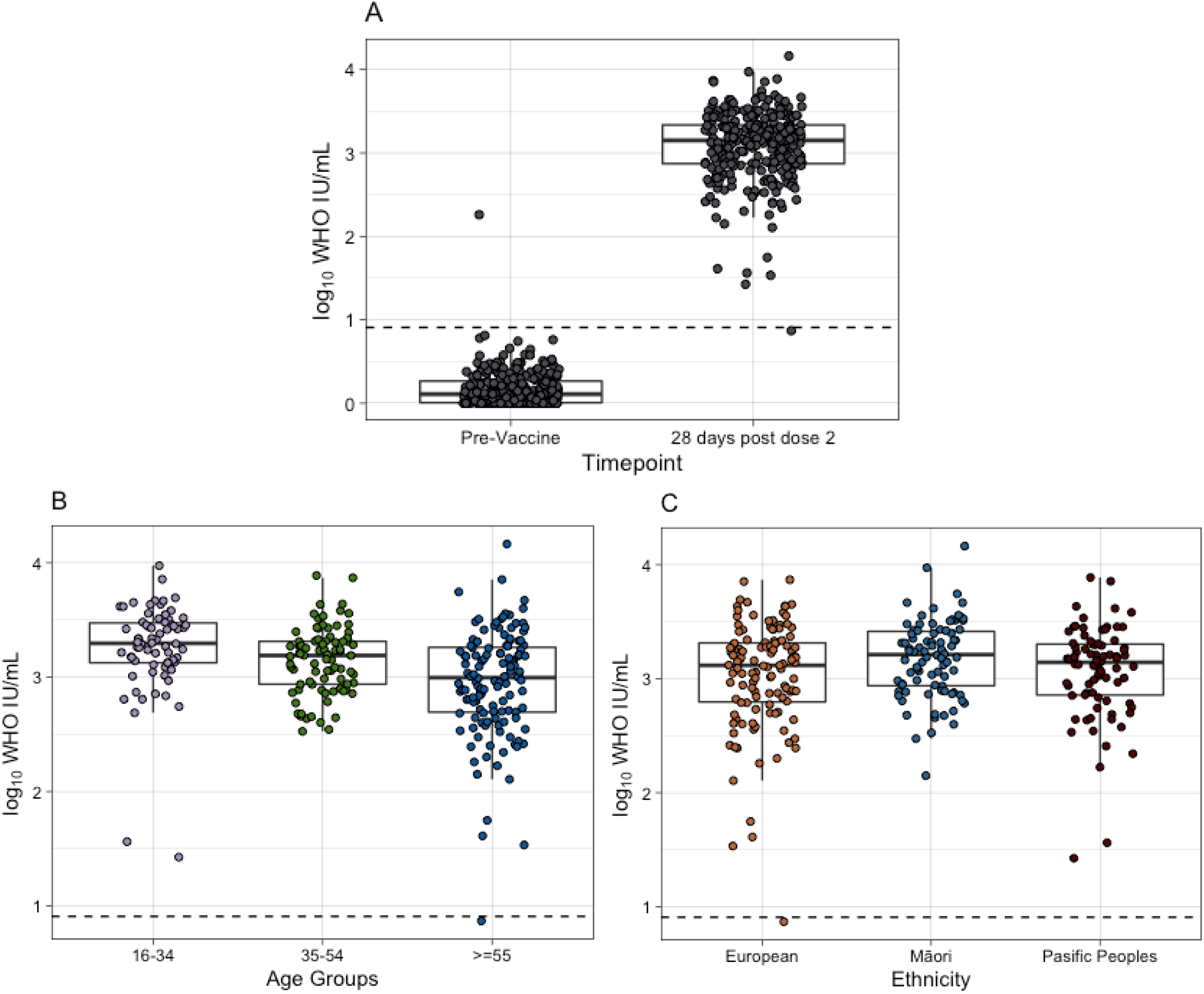
Unadjusted SARS-CoV-2 anti-S IgG responses by age and ethnicity (A) Unadjusted log_10_ transformed WHO IU SARS-CoV-2 anti-S IgG responses for participants pre-first vaccination and 28 days post-second vaccination, N = 285. (B) Data shown by age groupings, N=285. (C) Data shown by major ethnicity categories from NZ Census, N = 285. Solid horizontal line represents median and box represents interquartile range. The dotted line represents the log_10_ transformed value of the cut-off for a positive result (7.1 WHO IU/mL).

### Quantitative antibody response after vaccination

At 28 days after second vaccination, anti-S IgG antibody levels were high across gender and ethnic groups (Table 2, Figure 2b, Figure 2c). Geometric mean titers (WHO IU/mL) were similar between males and females. In an adjusted analysis controlling for age, ethnicity and BMI, GMT declined with increasing age: 1809 (95% CI 1446-2263) in participants 16-34 years of age, 1357 (95% CI 1089-1691) in participants 35-54 years of age, and 842 (95% CI 691-1027) in participants ≥55 years (p<0.001). Looking more closely at the ≥55 years of age group, GMTs did not differ significantly between 55-64 years of age and 65-74 years of age groups but were significantly lower in ≥75 years of age group (Table 3). GMT for participants ≥75 years of age was approximately half that of the 65-74 years age group (geometric mean ratio (GMR) 0.517) (p<0.001). In contrast, GMT for participants 16-34 years of age was significantly higher than other groups, with GMR of 1.35 compared to 35-54 years of age group, 1.58 compared to 55-64 years of age group, 2.0 compared to 65-74 years of age group, and 3.9 compared to ≥75 years of age group.

**Table 2.** SARS CoV-2 anti-S IgG and neutralising antibody responses after 2 doses of BNTb162b vaccine, adjusted for age, ethnicity and BMI

| Anti-S IgG |  |  |  |  | Neutralising antibody (ancestral strain) |  |  |  |
| --- | --- | --- | --- | --- | --- | --- | --- | --- |
|  | GMT<br>(AU/ml) | 95% CI | GMT<br>(WHO IU) | 95% CI | %<br>inhibition | 95% CI | GMT<br>(WHO IU) | 95% CI |
| **Gender** |  |  |  |  |  |  |  |  |
| Female | 8558 | 7258-10092 | 1215 | 1031-1433 | 94.4 | 93.7-95.0 | 1206 | 1044-1393 |
| Male | 9514 | 7929-11415 | 1351 | 1126-1621 | 94.5 | 93.8-95.2 | 1223 | 1043-1434 |
| **Age** |  |  |  |  |  |  |  |  |
| 16-34 | 12739 | 10184-15934 | 1809* | 1446 -<br>2263 | 95.3 | 94.5-96.0 | 1448† | 1191-1760 |
| 35-54 | 9556 | 7667-11910 | 1357* | 1089 -<br>1691 | 94.9 | 94.0-95.6 | 1341† | 1107-1625 |
| ≥55 | 5932 | 4868-7229 | 842* | 691 -<br>1027 | 93.0 | 91.9 | 919† | 773-1091 |
| **Ethnicity** |  |  |  |  |  |  |  |  |
| European | 9327 | 7736-11246 | 1325 | 1099 -<br>1597 | 94.4 | 93.6-95.1 | 1173 | 996-1381 |
| Māori | 10759 | 8662-13366 | 1528 | 1230 -<br>1898 | 95.1 | 94.3-95.8 | 1414 | 1170-1708 |
| Pacific<br>peoples | 7196** | 5694-9093 | 1022 | 809 -<br>1291 | 93.8 | 92.7-94.7 | 1076 | 877-1320 |
| **BMI** |  |  |  |  |  |  |  |  |
| ≤25 mg/m<sup>2</sup> | 9323 | 6853-12683 | 1324 | 973 -<br>1801 | 94.7 | 93.5-95.7 | 1309 | 1001-1712 |
| >25 kg/m<sup>2</sup> -<br><30 kg/m<sup>2</sup> | 7873 | 6434-9634 | 1118 | 914 -<br>1368 | 94.1 | 93.2-94.9 | 1121 | 940-1337 |
| ≥30 kg/m<sup>2</sup> | 9838 | 8463-11437 | 1397 | 1202 -<br>1624 | 94.5 | 93.9-95.1 | 1215 | 1066-1386 |
| **Diabetes** |  |  |  |  |  |  |  |  |
| Present |  |  | 770*** | 528 -<br>1121 | 91.3 | 88.8-93.3 | 710‡ | 513-981 |
| Absent |  |  | 1209 | 1053 -<br>1389 | 94.4 | 93.8-94.9 | 1188 | 1054-1338 |

\*p<0.001
\*\*p=0.025
\*\*\*p=0.022
†p=0.001
‡p=0.003

**Table 3.** SARS CoV-2 anti-S IgG responses after 2 doses of BNTb162b vaccine, adjusted for ethnicity and BMI

|  | % (n) | GMT<br>(AU/mL) | 95% CI |
| --- | --- | --- | --- |
| **Age** |  |  |  |
| 16-34 | 24.6 (70) | 12883* | 10383-15986 |
| 35-54 | 30.5 (87) | 9546 | 7720-11804 |
| 55-64 | 19.3 (55) | 8136 | 6300-10506 |
| 65-74 | 9.1 (26) | 6470 | 4520-9261 |
| ≥75 | 16.5 (47) | 3344** | 2465-4535 |

\*p<0.05 compared to each group
\*\*p<0.001 compared to each group

BMI was not significantly associated with anti-S IgG antibody. Pacific peoples were more likely to have lower GMTs (p=0.025). However, when controlling for the higher prevalence of diabetes in Pacific peoples subgroup (21%), GMTs did not differ by ethnicity. In an adjusted analysis of anti-S IgG antibody GMTs by comorbidity status, controlling for age, ethnicity and BMI, participants with diabetes had lower GMTs, 770 (95% CI 528-1121), compared to participants without diabetes, 1209 (95% CI 1053-1387) (p=0.022).

### Neutralising antibody response to ancestral strain (Wuhan) after vaccination

At baseline prior to vaccination, neutralising antibody responses to SARS-CoV-2 ancestral strain (Wuhan) were negative in all participants. At 28 days after second vaccination, almost every participant, 284 (99.6%) had neutralising responses to the ancestral strain on which the BNT162b2 vaccine is based (Figure 3). Neutralisation capacity against ancestral strain was consistently high, with mean 94.2% inhibition (95% CI 93.7–94.6). Similar to binding antibody, neutralising responses were robust and did not differ across gender, ethnicity or BMI groups, but were associated with age (Table 2, Figure 4). Converting to WHO IU/mL, neutralising titers declined with increasing age: 1448 (95% CI 1191-1760) in participants 16-34 years of age, 1341 (95% CI 1107-1625) in participants 35-54 years of age and 919 (95% CI 773-109) in participants ≥55 years (p=0.001). In an adjusted analysis of neutralising antibody GMTs by comorbidity status, controlling for age, ethnicity and BMI, participants with diabetes had lower neutralising GMTs, 710 (95% CI 513-981), compared to participants without diabetes, 1188 (95% CI 1054-1338) (p=0.003). Neutralising responses measured by sVNT correlated well with anti-S IgG antibody responses, correlation coefficient *r*^2^=0.795 (data not shown).

**Figure 3.**
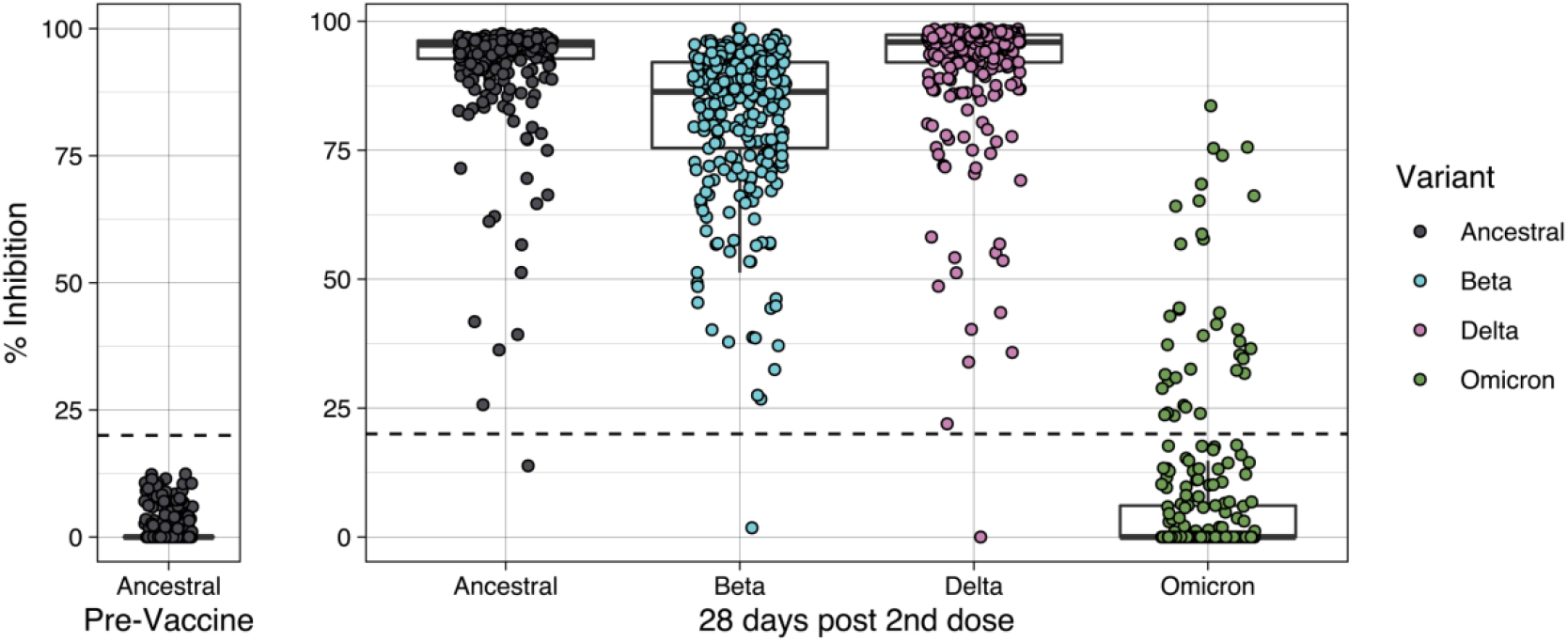
Unadjusted SARS-CoV-2 neutralising antibody responses to ancestral at baseline, and to ancestral and VoCs at 28 days post-vaccination Unadjusted percent inhibition against ancestral SARS-CoV-2 strain in participants pre-first vaccination (left pane) and inhibition against ancestral, Beta, Delta, and Omicron BA.1 variants from participants 28 days post-second vaccination (right pane), N = 285. Solid horizontal line represents median and box represents interquartile range. Cut-off for positive results is 20%.

**Figure 4.**
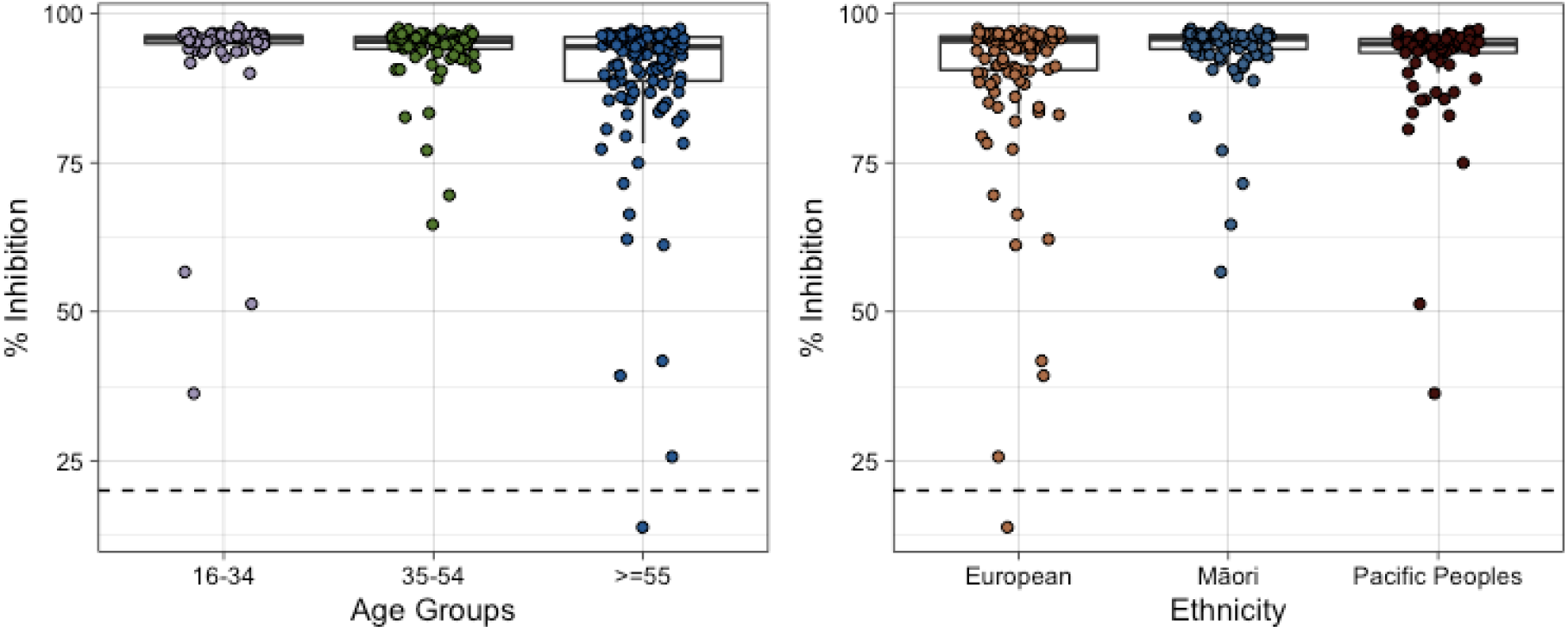
Unadjusted SARS-CoV-2 neutralising antibody responses to ancestral strain at 28 days post-vaccination by age and ethnicity Unadjusted percent inhibition against ancestral SARS-CoV-2 strain in participants post-second vaccination separated by age groups (left pane) and separated by ethnicity categories (right pane), N = 285. Solid horizontal line represents median and box represents interquartile range.

### Neutralising antibody response to Variants of Concern after vaccination

At 28 days after second vaccination almost every participant, 284 (99.6%) had antibodies that cross-neutralized Beta and Delta strains but only 36 (12.6%) had antibodies that cross-neutralized the Omicron BA.1 strain (Figure 3). Neutralisation capacity against Delta and the ancestral strain were high, with means of 94.9% and 94.2% respectively. Neutralisation of Beta was significantly lower than ancestral and Delta strains, mean inhibition of 86% (p<0.001). Neutralising responses to Omicron BA.1, where present, were 13-fold lower than responses to ancestral strain (p<0.001), with only 11 participants with >50% inhibition. Older age, but not ethnicity or BMI was associated with lower neutralising responses across VoCs (data not shown).

## DISCUSSION

Seroconversion rates, antibody response magnitude and neutralisation were consistently robust across gender, ethnicity, and most age groups after 2 doses of the BNT162b2 mRNA COVID-19 vaccine in this cohort of New Zealanders, enriched for older adults, Māori and Pacific peoples.

These findings are reassuring for New Zealand where the COVID-19 immunization strategy relied primarily on a single vaccine type across all populations.

This report represents one of the largest COVID-19 vaccine datasets on immune response in Māori and in people of Pacific Island background. It is reassuring to see no association of ethnicity on the humoral immune responses evaluated here. BMI also had no clear association with immune responses. Increased BMI has been associated with reduced immune response to other vaccines but not consistently [15,16]. Māori participants had high rates of pulmonary disease, primarily asthma, which may contribute to higher hospitalisation rates from COVID-19, but were not associated with reduced vaccine response. In contrast, Pacific participants were more likely to have diabetes, which was associated with reduced antibody responses.

As most IgG and neutralising assays used in the pivotal vaccine trials for COVID-19 vaccines were not standardized, it is difficult to compare across studies. In the pivotal trial of the BNT162b2 vaccine, IgG and neutralising antibody were evaluated with in-house assays in a population without previous SARS-CoV-2 infection that was 55% white and 0% Pacific [17]. At 28 days post-second dose, anti-S IgG titers were 7123 U/ml (95% CI 6217-8160) in ages 18-55 years, and 3961 (95% CI 3007-5217) in ages 56-85 years. Neutralising titers expressed as 50% neutralisation titers in a pseudovirus assay were 399 (95% CI 342-466) and 255 (95% CI 206-316) in ages 18-55 and 56-85 respectively. Real-world post-vaccination data using commercially available, validated assays are more readily comparable to the data in this study. Studies found very high seroconversion rates after 2 doses of BNT162b2 across age groups, similar to our findings [18,19]. Anti-S IgG titers were typically in the 3-4 log_10_ range (1000-10,000 U/mL), similar to our findings.

For example, a study in 483 primarily white and female healthcare workers with median age 41 in the UK used the Abbott Anti-S IgG Quant II assay and found a median titer of 10,058 AU/mL (95% CI 6408-15,582) in vaccinees without prior infection 1 month post-second dose of BNT162b2 [18]. A community-based study of 605 BNT162b2 recipients in the UK with median age of 60 used the Roche Anti-S IgG assay and found a median titer of 5311 U/mL (95% CI 3133-8829) a median of 42 days after second-vaccination [19]. Real-world data on neutralisation titer after 2 doses of BNT162b2 is typically reported using live viral or pseudoviral assays with titers in the 2-3 log_10_ range. For example, a study including 995 Israeli healthcare workers who received BNT162b2 vaccine found 50% neutralisation titer of 557 (95% CI 511-608) at 20 days post-second dose using a pseudoviral assay [20]. The sVNT assay used in this study has been shown to correlate with both pseudoviral and live viral neutralisation assays [21,22].

Both IgG and neutralising responses declined with age, with ≥75 years of age group significantly lower than other groups. This trend was found for BNT162b2 and other COVID-19 vaccines in both pivotal and real-world studies [17,23]. Lower immune responses to vaccination in older adults have been noted against several vaccine types and are generally attributed to immunosenescence and comorbidities. While IgG and neutralising titers in ≥75 year old age group were lower than other groups, they were still in the range associated with efficacy in pivotal trials and real-world studies of BNT162b2 and other mRNA vaccines [14,24–26]. Vaccine efficacy appears to increase with increasing IgG and neutralisation titer. In an analysis of a similar vaccine, mRNA-1723, vaccinees with 50% neutralisation titers of 10, 100 and 1000 at 4 weeks after the second dose had estimated vaccine efficacy of 78%, 91% and 96% [26]. However the ≥75 year old age group is at higher risk of antibody waning to sub-protective levels, as illustrated by more recent data showing waning antibody levels and decreased vaccine efficacy in older adults at 4-6 months after-second vaccination [6,27]. The current study findings support the relevance of booster vaccinations in NZ to restore waning immunity particularly in older adults.

Antibody levels were reduced in participants with diabetes, primarily type 2 diabetes. Diabetic participants were more likely to be Pacific peoples. While participants with diabetes were included in the pivotal trial of BNT162b2, limited data is available on antibody levels in this population. A previous report included 81 people with type 2 diabetes and found lower IgG and neutralising titers in this group [28]. In contrast, another report included a mixture of 150 people with type 1 and type 2 diabetes found no decline in IgG post-vaccination in diabetics [29]. However, vaccine efficacy appears to be maintained despite lower antibody responses. A large post-vaccination study in the US including 4931 healthcare workers estimated overall vaccine efficacy after BNT162b2 as 88.8% (95% CI 84.6-91.8) in diabetics compared to 80.2% (95% CI 45.8-92.7) in non-diabetics [30]. Follow-up data will be important to evaluate potential differences in waning of immune response in this group. If significant waning is demonstrated, an additional vaccine dose, similar to the three dose primary regimen plus booster currently used in immunosuppressed populations, could be assessed. This follow-up is also important from an equity perspective, given the higher rates of diabetes in Pacific peoples.

Consistent with international data, neutralisation capacity after 2 doses of BNT162b2 was severely reduced against the Omicron BA.1 variant with two-thirds of vaccinees having responses below the assay cutoff [5]. In contrast, the high % neutralisation titer was maintained against the Delta variant, with some decrease against the more neutralisation resistant Beta variant. Samples from the ongoing study will be assessed for VoC neutralisation after boosting, which has been shown to improve neutralising titers against Omicron BA.1 [5]. Future VoC can similarly be assessed to inform the level of protection in the NZ population going forward. Consistent with both international and local data, neutralisation correlated well with IgG antibody responses in this cohort [10,26].

This study has several limitations. It does not assess vaccine efficacy as there was little or no transmission during the study period described here. Only short term data through 28 days after 2 doses for the BNT162b2 vaccine is included. Understanding the durability of immune responses in this cohort, and response to booster doses will be important to assess the vaccination programme and shape future policy. Cellular immune responses are likely to be critical for both durability and breadth of COVID-19 vaccine-induced protection and are not included in the current study. Comorbidities were assessed by medical history and did not distinguish the duration or severity of disease, or whether the condition was treated/controlled. Cardiac and pulmonary comorbidities were grouped according to COVID-19 categories and include several diseases with varying pathology which may have diluted any potential associations with a single condition. This study did not include severely immunocompromised populations. Use of a high throughput surrogate viral neutralisation assay allowed rapid analysis for both ancestral and VoC responses, however there is less published comparative neutralisation data with this assay.

Vaccine antibody responses to the BNT162b2 were generally robust and consistent with international data in this COVID-19 naïve cohort with representation of key NZ and Pacific populations at risk for COVID-19 morbidity. Subsequent data on response to boosters, durability of responses and cellular immune responses should be assessed with attention to elderly adults and diabetics.

## Data Availability

All data produced in the present study are available upon reasonable request to the authors.

## Author contributions

FP, JU and GLG were responsible for study concept. FP and BL were responsible for study protocol. MW, SC, JM and GW were responsible for study conduct, data and sample collection. BL and KG were responsible for study coordination and data management. NM and RM were responsible for laboratory analysis. CF and BL were responsible for data analysis. FP, CF, BL and MB wrote the report. All authors reviewed and provided input to the report.

## Data availability

De-identified data can be made available upon request after review by MIMR.

## Acknowledgements

We would like to acknowledge the study participants and the study teams at both sites for their commitment to the study, including Michelle Sampson Lead Clinical Coordinator Lakeland Clinical Trials, Nina Linton Study Coordinator Lakeland Clinical Trials, Gary Lees Director of Nursing and Midwifery Lakes District Health Board, Ali Maoate Study Coordinator Southern Clinical Trials, Dr Monica Nua George Medical Director Etu Pasifika. Dr Api Talemaitoga provided input on study design and engaging Pacific peopless in the study. The Te Urungi Māori Advisory group of MIMR provided input on engaging Māori in the study. Mr John Treanor provided Māori consultation for Lakeland Clinical Trials site. Dr Matea Gillies was the Māori advisor, Ngai Tahu. Dr Peter McIntyre provided peer review of the study protocol for HDEC review. Giulia Giunti and Bethany Andrews at MIMR provided operational support for the study, and Aimee Paterson and Ciara Ramiah provided laboratory assistance at the University of Auckland.

## References

[1] Steyn N, Binny RN, Hannah K, Hendy SC, James A, Lustig A, et al. Māori and Pacific people in New Zealand have a higher risk of hospitalisation for COVID-19 | OPEN ACCESS. N Z Med J 2021;134:28–43.

[2] Koff WC, Schenkelberg T, Williams T, Baric RS, Adrian McDermott, Cameron CM, et al. Development and deployment of COVID-19 vaccines for those most vulnerable. Sci Transl Med 2021;13. https://doi.org/10.1126/scitranslmed.abd1525.

[3] COVID-19: Case demographics | Ministry of Health NZ n.d. https://www.health.govt.nz/covid-19-novel-coronavirus/covid-19-data-and-statistics/covid-19-case-demographics x(accessed March 29, 2022).

[4] Moore JP, Offit PA. SARS-CoV-2 Vaccines and the Growing Threat of Viral Variants. JAMA 2021;325:821–2. https://doi.org/10.1001/jama.2021.1114.

[5] Muik A, Lui BG, Wallisch AK, Bacher M, Mühl J, Reinholz J, et al. Neutralization of SARS-CoV-2 Omicron by BNT162b2 mRNA vaccine-elicited human sera. Science 2022;375:678–80. https://doi.org/10.1126/science.abn7591.

[6] Andrews N, Tessier E, Stowe J, Gower C, Kirsebom F, Simmons R, et al. Duration of Protection against Mild and Severe Disease by Covid-19 Vaccines. N Engl J Med 2022;386:340–50. https://doi.org/10.1056/NEJMoa2115481.

[7] Andrews N, Stowe J, Kirsebom F, Toffa S, Rickeard T, Gallagher E, et al. Covid-19 Vaccine Effectiveness against the Omicron (B.1.1.529) Variant. https://DoiOrg/101056/NEJMoa21194512022. https://doi.org/10.1056/NEJMoa2119451.

[8] Zimmermann P, Curtis N. The influence of the intestinal microbiome on vaccine responses. Vaccine 2018;36:4433–9. https://doi.org/10.1016/j.vaccine.2018.04.066.

[9] Craigie A, McGregor R, Whitcombe AL, Carlton L, Harte D, Sutherland M, et al. SARS-CoV-2 antibodies in the Southern Region of New Zealand, 2020. Pathology 2021;53:645–51. https://doi.org/10.1016/j.pathol.2021.04.001.

[10] McGregor R, Craigie A, Jack S, Upton A, Moreland NJ, Ussher J. The Persistence of Neutralising Antibodies up to 11 months after SARS-CoV-2 Infection in the Southern Region of New Zealand. N Z Med J 2021;135:162–6.

[11] Carlton LH, Chen T, Whitcombe AL, McGregor R, Scheurich G, Sheen CR, et al. Charting elimination in the pandemic: a SARS-CoV-2 serosurvey of blood donors in New Zealand. Epidemiol Infect 2021;149. https://doi.org/10.1017/S0950268821001643.

[12] Tan CW, Chia WN, Qin X, Liu P, Chen MIC, Tiu C, et al. A SARS-CoV-2 surrogate virus neutralization test based on antibody-mediated blockage of ACE2–spike protein–protein interaction. Nat Biotechnol 2020 389 2020;38:1073–8. https://doi.org/10.1038/s41587-020-0631-z.

[13] Whitcombe AL, McGregor R, Craigie A, James A, Charlewood R, Lorenz N, et al. Comprehensive analysis of SARS-CoV-2 antibody dynamics in New Zealand. Clin Transl Immunol 2021;10. https://doi.org/10.1002/cti2.1261.

[14] Zhu F, Althaus T, Tan CW, Costantini A, Chia WN, Van Vinh Chau N, et al. WHO international standard for SARS-CoV-2 antibodies to determine markers of protection. The Lancet Microbe 2022;3:e81–2. https://doi.org/10.1016/S2666-5247(21)00307-4.

[15] Frasca D, Blomberg BB. The Impact of Obesity and Metabolic Syndrome on Vaccination Success. Interdiscip Top Gerontol Geriatr 2020;43:86–97. https://doi.org/10.1159/000504440.

[16] Kageyama T, Ikeda K, Tanaka S, Taniguchi T, Igari H, Onouchi Y, et al. Antibody responses to BNT162b2 mRNA COVID-19 vaccine and their predictors among healthcare workers in a tertiary referral hospital in Japan. Clin Microbiol Infect 2021;27:1861.e1-1861.e5. https://doi.org/10.1016/j.cmi.2021.07.042.

[17] Public Assessment Report Authorisation for Temporary Supply COVID-19 mRNA Vaccine BNT162b2 (BNT162b2 RNA) concentrate for solution for injection. n.d.

[18] Eyre DW, Lumley SF, Wei J, Cox S, James T, Justice A, et al. Quantitative SARS-CoV-2 anti-spike responses to Pfizer-BioNTech and Oxford-AstraZeneca vaccines by previous infection status. Clin Microbiol Infect 2021;27:1516.e7-1516.e14. https://doi.org/10.1016/j.cmi.2021.05.041.

[19] Shrotri M, Navaratnam AMD, Nguyen V, Byrne T, Geismar C, Fragaszy E, et al. Spike-antibody waning after second dose of BNT162b2 or ChAdOx1. Lancet 2021;398:385–7. https://doi.org/10.1016/S0140-6736(21)01642-1.

[20] Levin EG, Lustig Y, Cohen C, Fluss R, Indenbaum V, Amit S, et al. Waning Immune Humoral Response to BNT162b2 Covid-19 Vaccine over 6 Months. N Engl J Med 2021;385:e84. https://doi.org/10.1056/NEJMoa2114583.

[21] Sholukh AM, Fiore-Gartland A, Ford ES, Miner MD, Hou YJ, Tse L V., et al. Evaluation of cell-based and surrogate SARS-CoV-2 neutralization assays. J Clin Microbiol 2021;59. https://doi.org/10.1128/JCM.00527-21.

[22] Murray MJ, McIntosh M, Atkinson C, Mahungu T, Wright E, Chatterton W, et al. Validation of a commercially available indirect assay for SARS-CoV-2 neutralising antibodies using a pseudotyped virus assay. J Infect 2021;82:170–7. https://doi.org/10.1016/j.jinf.2021.03.010.

[23] Koff WC, Williams MA. Covid-19 and Immunity in Aging Populations — A New Research Agenda. N Engl J Med 2020;383:804–5. https://doi.org/10.1056/NEJMp2006761.

[24] Khoury DS, Cromer D, Reynaldi A, Schlub TE, Wheatley AK, Juno JA, et al. Neutralizing antibody levels are highly predictive of immune protection from symptomatic SARS-CoV-2 infection. Nat Med 2021 277 2021;27:1205–11. https://doi.org/10.1038/s41591-021-01377-8.

[25] Bajema KL, Dahl RM, Prill MM, Meites E, Rodriguez-Barradas MC, Marconi VC, et al. Effectiveness of COVID-19 mRNA Vaccines Against COVID-19-Associated Hospitalization - Five Veterans Affairs Medical Centers, United States, February 1-August 6, 2021. MMWR Morb Mortal Wkly Rep 2021;70:1294–9. https://doi.org/10.15585/mmwr.mm7037e3.

[26] Gilbert PB, Montefiori DC, McDermott A, Fong Y, Benkeser D, Deng W, et al. Immune Correlates Analysis of the mRNA-1273 COVID-19 Vaccine Efficacy Trial. MedRxiv Prepr Serv Heal Sci 2021. https://doi.org/10.1101/2021.08.09.21261290.

[27] Nanduri S, Pilishvili T, Derado G, Soe MM, Dollard P, Wu H, et al. Effectiveness of Pfizer-BioNTech and Moderna Vaccines in Preventing SARS-CoV-2 Infection Among Nursing Home Residents Before and During Widespread Circulation of the SARS-CoV-2 B.1.617.2 (Delta) Variant — National Healthcare Safety Network, March 1–August 1, 2021. MMWR Morb Mortal Wkly Rep 2021;70:1163–6. https://doi.org/10.15585/MMWR.MM7034E3.

[28] Ali H, Alterki A, Sindhu S, Alahmad B, Hammad M, Al-Sabah S, et al. Robust Antibody Levels in Both Diabetic and Non-Diabetic Individuals After BNT162b2 mRNA COVID-19 Vaccination. Front Immunol 2021;12:4909. https://doi.org/10.3389/fimmu.2021.752233.

[29] Sourij C, Tripolt NJ, Aziz F, Aberer F, Forstner P, Obermayer AM, et al. Humoral immune response to COVID-19 vaccination in diabetes is age-dependent but independent of type of diabetes and glycaemic control: The prospective COVAC-DM cohort study. Diabetes Obes Metab 2022. https://doi.org/10.1111/dom.14643.

[30] Pilishvili T, Gierke R, Fleming-Dutra KE, Farrar JL, Mohr NM, Talan DA, et al. Effectiveness of mRNA Covid-19 Vaccine among U.S. Health Care Personnel. N Engl J Med 2021;385:e90. https://doi.org/10.1056/nejmoa2106599.

